# A Brief Acceptance and Commitment Therapy Intervention to Support Those Living At Risk of Inherited Prion Disease

**DOI:** 10.64898/2026.06.16.26355436

**Authors:** Rhianna Brien, Lilian Dindo, Rachel Williams, Lucy Pauli, Beth Marsh, John Collinge, Simon Mead, Edgar Chan

## Abstract

Living at risk of a neurodegenerative condition such as inherited prion disease (IPD) is associated with substantial psychological burden, yet evidence-based supportive interventions are lacking. This unmet need is likely to grow as advances in biomarkers and predictive testing lead to increasing identification of individuals in pre-symptomatic stages of neurodegenerative disease. Acceptance and Commitment Therapy (ACT), a transdiagnostic intervention targeting psychological flexibility, has shown promise in chronic health contexts but has not been evaluated in individuals at genetic risk. We conducted a feasibility and acceptability study of a brief, group-based ACT intervention in adults at risk of IPD recruited through the UK National Prion Clinic. The intervention comprised a single 5-hour, face-to-face workshop followed by an individual booster session. Prespecified feasibility and acceptability criteria were assessed alongside secondary psychological outcomes at baseline, 1 month, and 3 months post-intervention, complemented by semi-structured qualitative interviews. Twenty-three participants completed the intervention. All predefined feasibility criteria were met, including recruitment (58%), intervention completion (80%), retention at 3 months (79%), and low missing data (10%). Acceptability was high, with all participants reporting the intervention as useful and appropriate. Quantitative analyses demonstrated improvements in psychological quality of life and behavioural awareness at 3 months, with larger effects observed in participants with elevated baseline depressive symptoms. Qualitative findings highlighted the importance of peer connection, experiential learning, and practical strategies for managing uncertainty. These findings demonstrate that a brief, hybrid ACT intervention is feasible and acceptable for individuals living at risk of IPD and provide preliminary evidence for improving psychological well-being. As the population of individuals identified as at risk for neurodegenerative disease continues to expand, scalable psychological interventions that address cost and time barriers may represent an important component of future clinical care.

## Introduction

Despite growing recognition of the importance of psychological care within neurology, access to structured, evidence-based interventions remains limited, particularly for individuals living with risk rather than manifest disease. This is particularly pertinent as advances in biomarkers and predictive testing mean that increasing numbers of individuals will in future be living with knowledge of disease risk.

Prion diseases are rare, fatal neurodegenerative disorders caused by the accumulation of misfolded prion protein in the central nervous system. Inherited prion diseases (IPD) account for approximately 10-15% of cases and are autosomal dominant conditions caused by pathogenic PRNP mutations ^[1]^. They show wide variability in age of onset, symptom presentation, and rate of progression, even within families carrying the same mutation ^[2]^.

As there are currently no disease-modifying treatments, care primarily focuses on symptom management and maintaining quality of life ^[3]^. Psychological challenges associated with IPD often begin long before clinical onset, with elevated levels of anxiety and depression reported among at-risk individuals ^[4–5]^. As first-degree relatives have a 50% chance of carrying the pathogenic mutation, these individuals live with the ongoing uncertainty of whether they will develop a rare, incurable condition they likely have already witnessed in close relatives ^[5]^. Research in other inherited neurodegenerative conditions such as Huntington’s disease (HD) and familial Alzheimer’s disease has revealed similar psychosocial challenges; fluctuating distress shaped by individual circumstances and life events ^[6]^.

To date, no published research has examined psychological support for individuals at risk of IPD or prion diseases more broadly. Evidence from comparable conditions suggests that key needs may include managing emotional distress, coping with uncertainty about disease onset, and addressing legal and financial planning^[7]^. Preliminary studies have explored mindfulness-based interventions, psychoeducational forums, and relationship-focused approaches, with some evidence of reduced distress and greater peer support ^[6]^. However, even in HD, support for at-risk individuals remains scarce^[8]^. This likely reflects challenges including rarity, stigma, high attrition in longer interventions, and avoidance of psychologically distressing topics. Transdiagnostic, third-wave therapies such as ACT have been highlighted as promising for addressing these challenges^[9]^.

ACT has gained recognition as an alternative to Cognitive Behavioural Therapy (CBT), particularly for individuals facing chronic, uncontrollable stressors^[10]^. Rather than targeting symptom reduction, ACT aims to promote psychological flexibility, supporting engagement in meaningful, values-based actions in the presence of difficult thoughts and emotions ^[11]^ which may be particularly relevant for conditions like IPD that are characterized by persistent uncertainty^[10–12]^. Indeed, a growing body of research support the effectiveness of ACT across a range of chronic health conditions, including in neurological and neurodegenerative populations ^[10,12–16]^

However, existing ACT research in neurological populations has primarily focused on multi-session, individual formats. For individuals at risk of IPD, less intensive and more flexible approaches may be more feasible, given limited resources, geographical dispersion, and potential barriers to sustained engagement. Brief adaptable interventions may offer a more scalable and acceptable model of care in this context ^[17–18]^.

Single-session ACT interventions may be more feasible and acceptable than multi-session models^[19]^. So far, one-day ACT workshops have been implemented effectively across a range of medical and psychiatric conditions including diabetes, multiple sclerosis, and cardiovascular disease ^[20–22]^. Framing such interventions as “workshops” rather than “therapy” may also reduce stigma, better align with healthcare settings, and improve uptake^[23]^. In at-risk populations, group-based formats have also been associated with reduced isolation and stigma, as well as increased motivation and shared understanding^[6]^.

The present study aimed to investigate the feasibility and acceptability of a brief ACT-based intervention for individuals living at risk of IPD, comprising a single-session group workshop supplemented by an individual booster session. This hybrid format was designed to improve reach, accessibility, and engagements while maintaining opportunities for individualised support. Secondary aims examined changes in self-reported psychological measures.

## Method

Methods and reporting were guided by the CONSORT and TIDieR 2010 guidelines^24–26^.

### Participants and Procedure

Recruitment was conducted through the National Prion Clinic (NPC) and its National Prion Monitoring Cohort, a UK-wide longitudinal observational study established in 2008, and via advertising by the CJD Support Network. Participants were asymptomatic adults in the UK living at risk of IPD, either tested and positive for the gene mutation or untested. Exclusion criteria included active IPD symptoms, current suicidal ideation, severe mental health conditions, or significant cognitive impairment. All eligible participants were contacted by phone or email.

The study design was a non-randomised, parallel-group feasibility and acceptability trial with a waitlist control. An intended allocation ratio of 1:1 was used. Participants were allocated to either the intervention group (receiving the ACT workshop) or a waitlist control group (treatment as usual), based on availability to attend. Randomization was not feasible due to logistical constraints related to participant availability and individual needs, and maximizing participation was prioritised.

Outcome measures were completed online at three time points: two weeks pre-workshop (T1), four weeks post-workshop (T2), and three months post-workshop (T3).

All control group participants were offered to participate in the intervention following the study phase. After the intervention, control participants completed follow-up measures at T2 and T3.

### ACT Intervention

The intervention was a five-hour, face-to-face group workshop held at the NPC in London. Content was adapted from a single-session ACT protocol developed for other clinical populations by L.D. ^[27–28]^. Two focus groups of individuals at risk of IPD (n=5 & 2) were conducted during the study preparation phase to refine workshop content and participant materials. A pilot session was also conducted with staff members of the NPC, who collectively have had decades of experience working with patients and families with IPD, for further feedback and refinement.

The workshop followed a structured agenda covering the six core processes of ACT incorporating didactic teaching, experiential exercises, and group reflection to enhance psychological flexibility^[11]^. Core components were covered in each workshop, with some flexibility to adapt to the needs of each group. Participants were provided with a manual to take home. Four weeks post-workshop. participants received an individualized one-hour booster session delivered either online or by phone, to reinforce skills learned, identify barriers to implementation, and encourage ongoing practice.

The original workshop included some psychoeducation ^[27–28]^ but focus group feedback suggested IPD-specific content during the workshop could be confronting and detract from the primary aims. Accordingly, it was adapted into a 30-minute online video delivered by a clinical nurse specialist (R.W.), and sent to participants one week before the workshop.

All workshops were facilitated by a consultant clinical neuropsychologist (E.C.) and a trainee clinical psychologist (R.B.). Facilitators achieved ACT Level 1 accreditation and completed a two-day training course on delivering ACT in single-session workshop form by L.D. All workshops were audio-recorded, and adherence was independently assessed by an independent reviewer with a background in psychiatry (L.P.) using a pre-defined checklist of the six core ACT processes.

### Outcome measures

A mixed-methods approach was used to integrate quantitative and qualitative data. A priori feasibility criteria were established to guide progression decisions. These were: (a) ≥50% of contacted individuals consent to participate, (b) ≥70% retention at follow-up, (c) ≤10% missing data and (d) ≥80% intervention completion rate. Acceptability was measured using an adapted version of the Theoretical Framework of Acceptability (TFA) questionnaire post-workshop^29^. The acceptability criterion was that ≥70% of intervention participants would have a median score of ≥4 on the TFA. Qualitative feedback was also collected three months post-intervention via structured telephone interviews or written open-ended responses. Practically meaningful thresholds were chosen as per published guidance^30–31^.

Secondary outcomes were chosen to measure the potential effectiveness of the intervention. Based on previous findings from single-session ACT interventions ^[19]^, self-report measures focused on psychological distress, psychological flexibility, and quality of life. They included: 1) Depression Anxiety Stress Scales-21 (DASS-21), 2) Comprehensive Assessment of Acceptance and Commitment Therapy Processes (CompACT), and 3) World Health Organization Quality of Life-BREF (WHOQOL-BREF).

### Statistical Analyses

Feasibility outcomes were assessed in accordance with the original study design, with data considered separately by study arm to reflect procedures in a future randomized controlled trial. For acceptability and secondary outcomes, analyses were conducted on all participants who completed the intervention (i.e. intervention plus control participants). This approach maximised the available sample and enabled a more comprehensive evaluation of the intervention.

Feasibility and TFA data were analysed descriptively and narratively. Qualitative data were analysed using inductive thematic analysis, a flexible, data-driven approach ^[32]^.

Repeated measures ANOVAs were conducted on DASS-21, CompACT, and WHOQOL-BREF total and subscale scores across the three time points (T1, T2, T3). Missing data at T3 were handled using the Last Observation Carried Forward (LOCF) method. Given the small sample size and the exploratory nature of this pilot study, corrections for multiple comparisons were not performed, as these would have been overly conservative and risked overlooking potentially meaningful patterns. Analyses were interpreted cautiously with a focus on effect sizes and patterns rather than statistical significance alone

## Results

### Participant Characteristics

A total of 29 participants (10 men and 19 women), with a mean age of 45 years (range: 20-72) were allocated to a study condition: 15 to the workshop arm and 14 to the waitlist control (see Figure 1). Of these, 23 participants completed the intervention across 5 workshops, including 13 from the workshop arm and 10 from the waitlist control. The patient characteristics of those who completed the intervention is shown in Table 1. A range of gene variants was represented in the cohort, with E200K and P102L being the most common. Sixteen participants (70%) had tested positive for the prion gene variant, while the rest were untested. Most participants identified as White British; one identified as “Other Mixed” ethnic background. Ten patients (43%) had engaged in psychotherapy before attending the workshop, but only one had experience with ACT. Ten patients (43%) reported watching the pre-workshop psychoeducation video about IPD. All workshops demonstrated adherence to the pre-developed ACT protocol checklist, as evaluated by an independent reviewer (L.P.).

**Figure 1.**
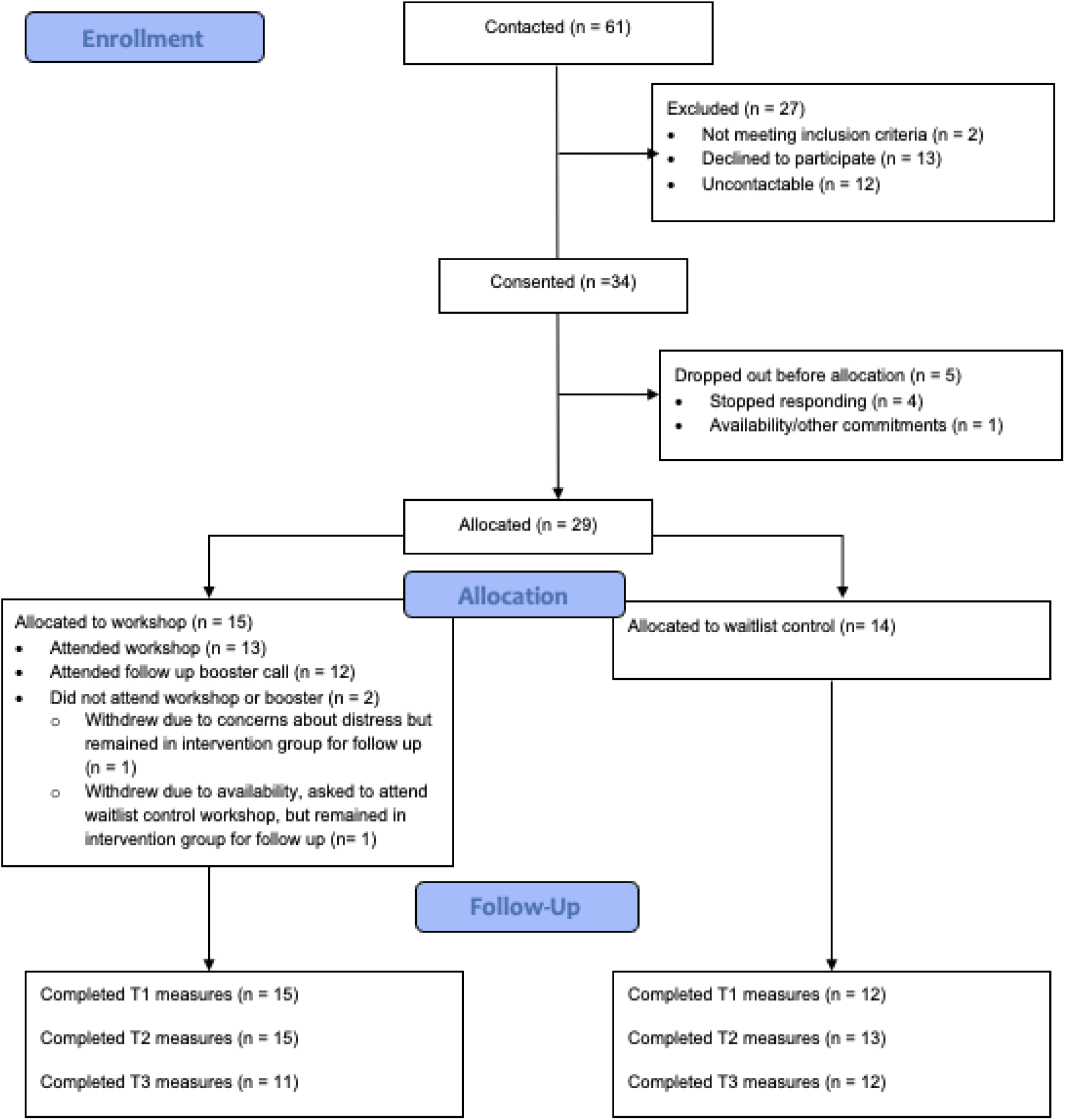
CONSORT flow diagram of participant recruitment, allocation and follow-up.

**Table 1.** Participant characteristics of those who received the intervention.

| Workshop | Gene variant | Gender | Testing status |
| --- | --- | --- | --- |
| 1 | E200K | Male | Untested |
|  | E200K | Female | Tested+ |
|  | E200K | Male | Tested+ |
|  | P102L | Female | Tested+ |
|  | D178N | Female | Tested+ |
| 2 | E200K | Female | Tested+ |
|  | P102L | Male | Untested |
|  | 6-OPRI | Female | Tested+ |
| 3 | E200K | Female | Tested+ |
|  | E200K | Female | Tested+ |
|  | E200K | Male | Tested+ |
|  | P102L | Male | Tested+ |
|  | P102L | Female | Untested |
| 4* | E200K | Female | Untested |
|  | E200K | Female | Untested |
|  | P102L | Female | Untested |
|  | D178N | Male | Tested+ |
|  | D178N | Female | Tested+ |
|  | Q212P | Female | Tested+ |
|  | 5-OPRI | Female | Untested |
| 5* | E200K | Female | Tested+ |
|  | E200K | Female | Tested+ |
|  | E146G | Female | Tested+ |

*\*Workshops were conducted after the study phase with participants from the control arm*

### Feasibility

Of the 61 individuals contacted, 59 were eligible and 34 consented, yielding a recruitment rate of 58% (see Figure 1). Reasons for ineligibility included recent symptom onset and cognitive comorbidities. Among those eligible but not recruited 13 declined participation – citing schedule conflicts, lack of perceived need for the intervention, or concerns about burden – and 12 could not be contacted.

Five participants withdrew after consenting but before allocation, resulting in 29 participants (85%) assigned to study arms. Of the 15 participants assigned to the workshop arm, 13 completed the workshop and 12 completed the booster call yielding an overall completion rate of 80% based on the 15 originally assigned.

Retention rates exceeded the *a priori* target of ≥70% at follow-up, with measure completion rates of 93% at T1, 97% at T2, and 79% at T3. Retention was calculated at each time point based on the original allocated sample (N = 29).

Across all time points (T1–T3), nine out of 87 expected outcome measure sets (29 participants × three time points) were not completed, resulting in an overall missing data rate of 10%. Mean completion times for the outcome measures indicated low participant burden and supported their feasibility: 2.7 minutes for the DASS-21 (SD = 1.4, range = 0.9–7.5), 5.5 minutes for the CompACT (SD = 4.3, range = 1.1–36.2), and 3.6 minutes for the WHOQOL-BREF (SD = 2.1, range = 1.2–12.0).

### Acceptability

The a priori acceptability criterion was met. Specifically, 21 out of 23 participants (91%) had a median score of 4 or above on the TFA when scores were aggregated across all 9 items. All 23 participants rated the intervention as acceptable or completely acceptable. Across the 9 items, the most common responses were positive: participants liked it, found it useful, understood how it worked, and did not feel it interfered with other life priorities. There was more ambivalence around effort and confidence. Full response distributions are presented in Figure 2a.

**Figure 2.**
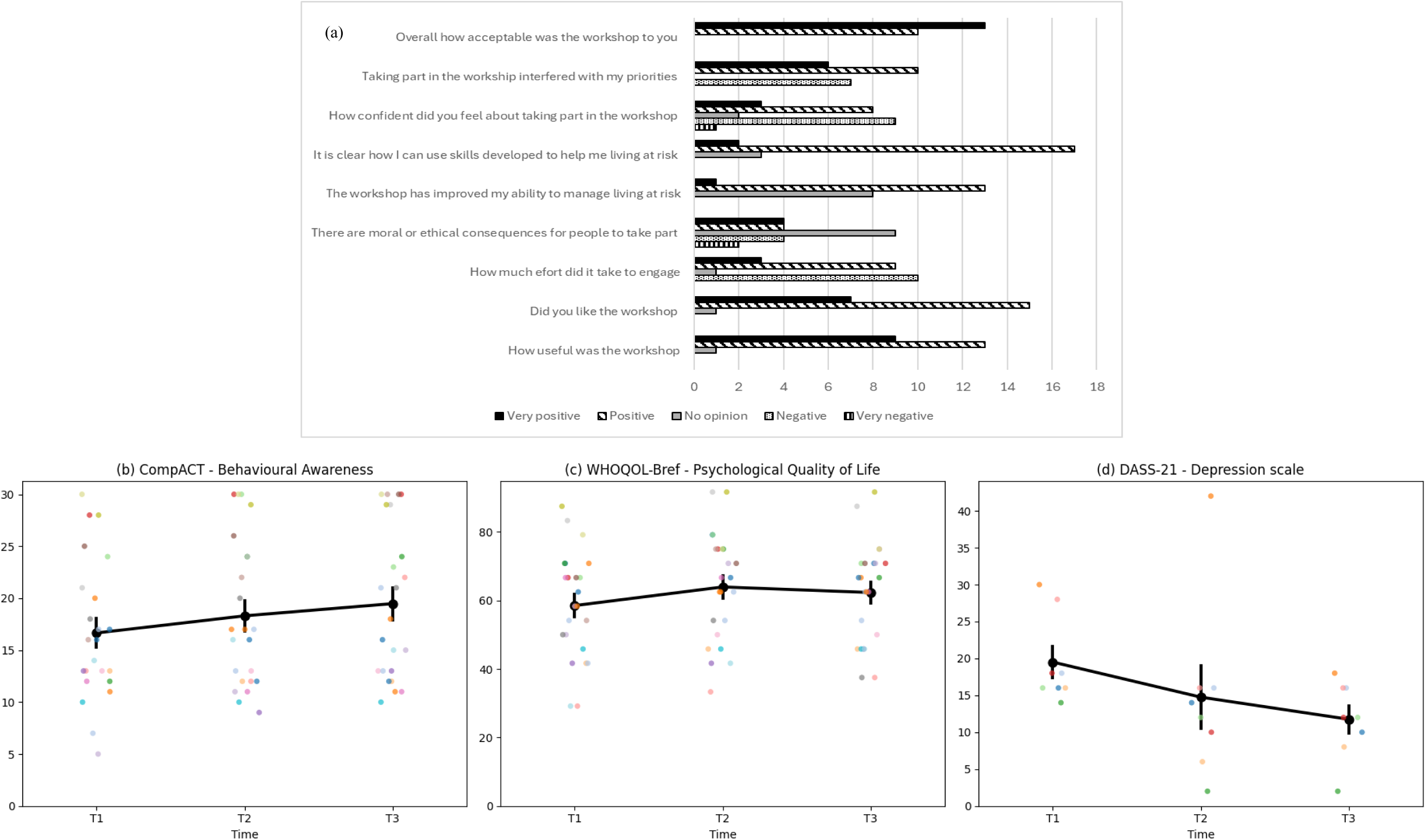
Acceptability responses and secondary outcomes. (a) Responses to the Theoretical Framework of Acceptability questionnaire. Response options were tailored to each item but are displayed here in a generic format (very positive to very negative) for ease of interpretation. Mean response score and standard error on the (b) CompACT Behavioural awareness scale and (c) WHOQOL-Bref Psychological quality of life, across the three time points. Higher scores reflect better psychological functioning. (d) Mean response score and standard error on the DASS-21 Depression scale across the three time points for those who reported moderate and above depression symptoms at T1. Lower scores reflect less severe depressive symptoms.

### Thematic Analysis of Participant Feedback

Fourteen participants (61%) who completed the workshop provided qualitative feedback post workshop either through structured telephone interviews or open-ended response forms. Five themes relevant to intervention acceptability were developed through thematic analysis. Selected participant quotes are shown in Table 2.

**Table 2.** Themes and Illustrative Quotes from Participant Interviews.

| Themes | Subthemes | Example quotes |
| --- | --- | --- |
| **1. Structure and delivery** | 1.1 Session length and timing | “It was for the right amount of time,” “Positive that it was one block,” “about right”. |
|  | 1.2 Group size and interaction | “Felt safe to chat and express how we felt”, “A balance between intimate space and more people.”, “It was a nice small group so didn’t feel overwhelming,” “the small group meant that people were quite open.” |
|  | 1.3 Facilitation experience | “Both facilitators were very good, relaxed and funny and made it interesting at a level to suit us all.” |
| **2. Workshop experience** | 2.1 Social connection | “A good opportunity to meet others” (Martin) “The girl sitting next to me said, well her life has changed so she may want to rethink that... and I hadn’t even thought about that” (Caroline) |
|  | 2.2 Expectations versus reality | “I was expecting to have more chat around the condition... to speak with those other members about how we felt and lived with the condition.”, “I was not 100% sure what to expect, but it met with what I hoped it would cover.” |
|  | 2.3 Engagement with ACT | “Gives you the opportunity to look at what your barriers are and maybe change them”, “Identify the difference between carrying something...but not being consumed by it.”, “writing down a goal and how to get there was useful,” |
| **3 Usefulness and relevance** | 3.1 Perceived value and resonance | “Useful as it can provide ways to deal with difficult feelings and emotions that can come up around it.”, “Some parts were very helpful, some parts I never really understood” |
|  | 3.2 Practical tools and takeaways | “It has given me some strategies”, “Good for everyday life” |
|  | 3.3 Follow up call and materials | “Good to catch up...and work though some of my thoughts after.”, “I haven’t used it since attending...but it is a good thing to have.”, “Very clear, descriptive, and informative but easy to follow. I’ve used it since and would revisit it if I had struggles.” |
| **4 Improvements for the future** | 4.1 Depends on stage of journey | “I think it would be more useful for people who are recently coming to the diagnosis.”. “I’d quite like to do it again further down the line.”. |
|  | 4.2 Suggested refinements | “More check in sessions / booster calls over a longer period of time” |
| **5. Condition-Specific Needs and Expectations** | 5.1 Balance of content | “I think if they did make reference to the reason why we were there...that would be powerful” |

#### Theme One: Structure and Delivery

Most participants found the workshop length and structure appropriate. Minor suggestions included slight adjustments to duration, such as shortening the day slightly, adding an additional hour to allow more to be covered, or allowing more time for breaks. The small group format was consistently valued, described as intimate, safe, and conducive to sharing. Facilitation was viewed positively, with a relaxed and engaging style supporting discussion of difficult topics.

#### Theme Two: Workshop Experience

Participants repeatedly highlighted the value of social connection and shared experience. Hearing from others “in the same sort of situation” or with “similar experiences” was described as supportive and at times “quite powerful.” Many valued the opportunity to speak openly, recognising that such conversations were often difficult to have with friends or family.

Expectations varied, with some anticipating more disease-specific discussion and feeling “unsure” about whether they were “supposed” to raise this, while others were satisfied with the content. Engagement with ACT components was high, with techniques such as breathing exercises, noticing thoughts, values clarification, and goal-setting reported as particularly helpful. Several reflected on key learnings such as “letting thoughts come and go” and “focusing on the more important things.” Several described finding experiential tasks “powerful” in shifting perspectives or helping them approach things differently.

#### Theme Three: Usefulness and Relevance

Most participants reflected on finding the workshop useful. Many reported gaining “strategies,” “life skills,” or new ways to manage stress and difficult feelings. Some described significant benefits, including feeling better able to cope, reduced stress, or going days without thinking about prion disease – something they had not previously experienced.

The follow-up call was consistently valued for offering an opportunity to reflect in a more personal and focused way. Participants appreciated being able to explore topics they did not raise in the group and felt that the conversation helped consolidate learning. Several described the call as “thorough,” “fulfilling,” or more tailored and personal than the workshop.

Use of the printed materials was mixed. Some participants had re-read or used the handbook, while others had not gone through it since the workshop but appreciated having it. A few participants noted keeping it available at home and finding reassurance in knowing it could be revisited when needed.

#### Theme Four: Improvements and Tailoring

Participants reflected the workshop was particularly relevant for those newly coming to terms with a genetic diagnosis or family history. Others believed it might be beneficial to revisit the content at different life stages.

Suggestions for refinement included extending break times, offering more opportunities for group bonding (e.g., longer lunches or morning introductions), and incorporating additional booster calls or check-ins. Some participants emphasised the value of face-to-face follow-ups, while others wanted more unstructured time to build rapport with the group.

#### Theme Five: Condition-Specific Needs and Expectations

A significant proportion of participants expressed a desire for more explicit discussion on prion disease and experiences of living at risk. Several participants also expressed interest in receiving more factual or clinical information such as what is currently known about the condition, recent advances in research, and questions about gene expression and epigenetics.

Finally, group composition was important for some attendees. Several suggested that grouping participants by risk status or shared lived experience (e.g., gene-positive, untested, affected gene type), could enhance empathy, relevance, and openness.

### Secondary Outcomes

Repeated measures ANOVAs were conducted to evaluate the impact of the intervention on DASS-21, CompACT, and WHOQOL-BREF total and subscale scores across the three time points. Analyses revealed significant improvement in the Behavioural Awareness subscale of the CompACT (F(2, 44) = 5.22, p=0.01, η2p=.19) which measures present-moment awareness and mindfulness in daily life. Pairwise comparison revealed significant difference between T1 and T3 (p=0.01), but not between T1 and T2 (p=0.05) or between T2 and T3 (p=0.15). There was also significant improvement found on the psychological quality of life domain from the WHOQOL-BREF (F(2, 44) = 7.08, p=0.00, η2p=.24; see Figure 2b,c) which measures mental and emotional well-being. Pairwise comparison revealed significant difference between T1 and T2 (p=0.00), T1 and T3 (p=0.04) and not T2 and T3 (p=0.13). All other variables had p>0.1 except Total CompACT score (F(2,44)=3.06, p=0.06, η2p=.12).

As this was a pilot study, all interested and eligible individuals were included to maximise participation, regardless of psychological well-being. However, this may have obscured effects among participants with elevated distress, so secondary analyses were conducted for only those reporting moderate or greater DASS-21 symptoms at T1 (n=8).In this subgroup, Depression scores improved significantly (F(2,14) = 4.77, p = 0.03, η²ₚ = 0.41; see Figure 2d), with significant pairwise difference between T1 and T3 (p=0.00). CompACT total scores also improved (F(2,14) = 5.87, p = 0.01, η²ₚ = 0.46), as did the social domain of the WHOQOL-Bref (F(2,14) = 3.92, p = 0.04, η²ₚ = 0.36), reflecting gains in satisfaction and functioning in social relationships and support networks. Consistent with the whole sample, behavioural awareness and psychological quality of life also showed significant improvements, with larger effect sizes observed in this subgroup (BA: F(2,14) = 6.24, p = 0.01, η²ₚ = 0.47; Psych: F(2,14) = 15.85, p<0.00, η²ₚ = 0.69).

## Discussion

We demonstrated that a brief ACT intervention comprising a five-hour, in-person group workshop followed by an individual booster session was both feasible and acceptable for individuals living at risk of IPD. Secondary outcomes suggested positive psychological benefits at 3 months follow-up, particularly for individuals who were experiencing moderate to severe depressive symptoms prior to the intervention. These findings provide preliminary support for this hybrid-format ACT approach in this population.

Our study met all four predefined feasibility targets, with recruitment at 58%. Participants who declined or withdrew most reported that they did not require the intervention or were concerned that participation might negatively affect them. For those who engaged with the intervention, completion and retention were high. This suggests that the intervention and outcome measures were not overly burdensome, supporting feasibility in future studies.

All participants rated the intervention as acceptable. Most participants liked it, found it useful, and understood how it worked and could be applied to their lives. There was greater ambivalence in responses relating to effort and confidence, perhaps reflecting uncertainty regarding what the workshop might entail.

Qualitative feedback indicated that both the length and content of the workshop were acceptable, relevant and generally well received. The skills and strategies introduced were viewed as useful and applicable to managing uncertainty and emotional challenges associated with being at risk of neurological disease. Peer interaction was described as particularly beneficial, echoing findings from interventions targeted at other at-risk populations such as HD ^[6]^.

Feedback regarding the extent of discussion about prion disease during the workshops was mixed. Some participants felt the balance was appropriate, whereas others suggested that the topic could have been addressed more explicitly, consistent with previous research which suggests the importance of tailoring psychological interventions to the condition, context and an individual’s readiness to engage with open discussion ^[18]^. Some participants also reflected they preferred to be grouped with others of similar testing status or gene variant, reflecting the heterogeneity within this population ^[2]^. In the pilot groups, participants indicated that more direct discussion of prion disease could feel unhelpful or overly confronting, particularly given the varied experiences and stages of adjustment among individuals attending the groups. Thus, it was decided that mention of prion disease would be mainly participant-led rather than explicitly integrated within the workshop content. Feedback from those who have completed the intervention suggests that future iterations of the programme should clarify in advance the expected level of disease-specific discussion. It may also be helpful to offer participants the option of attending groups with differing emphases with separate groups focused primarily on psychological coping strategies versus those allowing more open discussion of disease-specific concerns.

Secondary outcomes showed that following the intervention, participants experienced improved behavioural awareness and psychological quality of life. The improvement was significant 4 weeks after the workshop and sustained at 3 months. In those with elevated depression scores pre-intervention, depression scores significantly declined at 3 months, accompanied by improvements in psychological flexibility, social and psychological quality of life. Our findings add to the growing evidence-base that the transdiagnostic ACT framework is well-suited to neurological and neurodegenerative diseases ^[10, 15–16, 22]^, particularly through its focus on acceptance, values-based action, and coping with difficult emotions.

Our positive findings also suggest that a single-session group format is a practical and effective method of delivering ACT. All those who attended the workshop stayed for the entire duration, covering all the key concepts of ACT which would normally be covered across 4-6 separate 1-hour sessions. A single-session format reduces attrition and increases the likelihood that patients receive the full course of therapy. In the UK, it has been shown that the modal number of sessions attended for weekly CBT therapy is 2, with 33% of patients referred to high intensity treatment attending only 1 session^[33]^. In HD, attrition rates is often related to ambivalence about engagement and possible avoidance of psychological distress ^[8]^. A single-session workshop format also has practical advantages such a reducing travel, time commitment, administration and overall costs. The inclusion of an individual booster session may have helped consolidate skills and support maintenance of gains over time, highlighting its potential value for enhancing longer-term impact.

Several limitations should be noted. The small sample meant secondary outcomes were analysed in all intervention completers, limiting comparison with treatment as usual. However, improvements in ACT-specific measures suggest effects are unlikely solely due to non-specific factors. Future trials should incorporate an appropriate comparator condition. Given that participants highlighted the value of peer support within the workshop, it may be particularly informative to include an active control condition, such as a group-based peer support session, to better isolate the specific effects of the intervention.

The sample was drawn mainly from a highly engaged research cohort, limiting generalisability to less engaged or under-served individuals who may have different needs or distress levels. This may also have influenced recruitment rates, an important consideration for future full trials given the rarity of IPD. The sample also included a higher proportion of tested individuals (69%) than would be expected based on broader population estimates, which may further skew findings. Evidence suggests that those who undergo testing tend to cope better psychologically, potentially inflating perceived acceptability or benefit^[34]^.

This is the first study to trial a psychological intervention to support individuals at risk of IPD. Findings support the feasibility and acceptability of a brief ACT-based intervention and provide preliminary evidence that it can improve psychological well-being. The hybrid format may offer a practical, resource-efficient model for supporting people at risk of rare conditions where specialist access is limited, with potential applicability to other neurological conditions. The brief format may be particularly advantageous within constrained healthcare systems, where access to extended psychological therapy is limited.

## Data availability

The data that support the findings of this study are available from the corresponding author upon reasonable request. Restrictions apply to the availability of these data due to their sensitive nature and to preserve participant confidentiality.

## Acknowledgement

We thank Elena Binns and Rowena Baker for their assistance in the administration of the project, and all the staff at the National Prion Clinic and the CJD Support Network for their support.

## Patient consent

Ethical approval for this study was obtained as an amendment to the broader NPMC study. (Ref: 05/MRE/0063). All participants provided written informed consent prior to participation.

